# Citizen Science and Public Health- Can eBird data inform relationships between public health and access to biodiversity?

**DOI:** 10.1101/2022.01.04.22268764

**Authors:** J.J. LaFantasie

## Abstract

The relationship between access to nature and public health outcomes has been well-studied and established in the literature. However, most studies use simple greenness indices as a proxy for access to nature, which ignores the “quality” of the nature since greenness indices are not able to predict biodiversity. My objective was to investigate the relationship between citizen scientist collected biodiversity data from the eBird platform, urban greenness and four human health outcomes (asthma, coronary heart disease, and self-assessed mental and physical health). I mapped and tested for correlations among eBird record species richness, greenness as NDVI and PLACES human health data in urban census tracts located in three metro areas/ecological zones (Albany, NY: eastern deciduous forest, Kansas City, MO: tallgrass prairie, and Phoenix, AZ: Sonoran Desert). eBird species richness was related to greenness, measures of urbanization and several human health factors; however, the correlations varied by metro area and in strength. Provided confounders are controlled for, eBird data could help to refine models surrounding relationships between public health and nature access.

---

It is established in the literature that proximity and access to “nature” and green space is related to subjective wellness and health outcomes (Jennings et al. 2016, Mavoa et al. 2019). Greenness indices are often used to evaluate the availability of natural areas in urban landscapes, and while NDVI is often a good estimator of green spaces, in some situations it may be over-simplification to suggest that the health benefits of nature areas are simply a function of local greenness (Markevych et al. 2017, Reid et al. 2018, Jarvis et al. 2020). There is also evidence that exposure to biodiversity, that is, the “quality” of the green space and the variety of habitats it offers, influences health outcomes (Fuller et al. 2007, Hanski et al. 2012, Shanahan et al. 2015, Mills et al. 2020). It is often assumed that greenness and biodiversity are synonymous and yet a functional crop field can be low in biodiversity but have a high NDVI. While there have been many attempts to model diversity based on vegetative reflectance, effectiveness and portability of these methods vary (Wang and Gamon 2019, Leveau et al. 2020), and ground truthing biodiversity is costly. Investigation of the mechanisms behind the relationship between greenness and health outcomes continues (Markevych et al. 2017); this study proposes to further probe this question using citizen gathered data from the eBird platform (Cornell Lab of Ornithology 2020), particularly in urban areas.

With rapid increases in data and data-driven decisions, citizen science has begun to play a larger role in scientific investigations, despite its inherent flaws. eBird, which was initiated in 2002, is a well-established, crowdsourced, georeferenced database to which more than 600,000 citizens and scientists submit bird sightings. The quantity of eBird data collected by citizen birders may be useful in further elucidating relationships among greenness, biodiversity, and human health. Urban development tends to homogenize bird communities, meaning that species richness and diversity decrease at high urban densities (Melles 2005, Tratalos et al. 2007), and trends in bird diversity and density may indicate biodiversity status in general. I may be able to use eBird contributor record species richness to provide a proxy measure of biodiversity and relate this to greenness and human health.

My objectives were to: 1) Map eBird records of bird species and determine bird species richness at the census tract level, and 2) Relate eBird record species richness to census tract level NDVI, development intensity and local human health outcome measures to elucidate relationships.

## Methods

I mapped eBird, greenness as NDVI and CDC PLACES human health data in urban census tracts located in three metro areas (Albany, NY: eastern deciduous forest, Kansas City, MO: tallgrass prairie, and Phoenix, AZ: Sonoran Desert). I purposely located the areas in three distinct ecological zones to capture variation in response due to bird species distributions and climate and to avoid potentially faulty conclusions that can occur when using a single site (Fuertes et al. 2014). Temporally, I limited my analyses to only 2018 eBird sightings and NDVI data; I chose 2018 because the population density (Manson et al. 2020) and CDC public health PLACES data are dated to 2018.

### Data processing/analysis

I downloaded census, public health, satellite reflectance, eBird and land use data (Table 1). For each metro area, I selected urban tracts from census data (Manson et al. 2020) based on calculated population density > 1500 people/square mile and at least 40% developed area based on USGS-NLCD 2011 land cover rasters (2014). I calculated the number of unique bird species sighted and mean NDVI values for each tract and spatially joined these and the PLACES health data to the urban tract polygons (USGS 2018, Centers for Disease Control et al. 2020). I calculated bivariate Spearman’s correlation coefficients to determine associations among eBird record species richness, NDVI, and public health outcomes,. I focused on four health outcomes (asthma, coronary heart disease and self-rated mental and physical health based on prior work (Hanski et al. 2012, James et al. 2015) and the self-rating aspect of the general mental health and physical health fields. I also calculated and visualized relationships among eBird species richness, NDVI and population density using the same methods. I used ArcGIS 10.6.1 and R for all data analysis and visualization (Buckley 2015, Esri 2017, R. Core Team 2020).

**Table 1.**
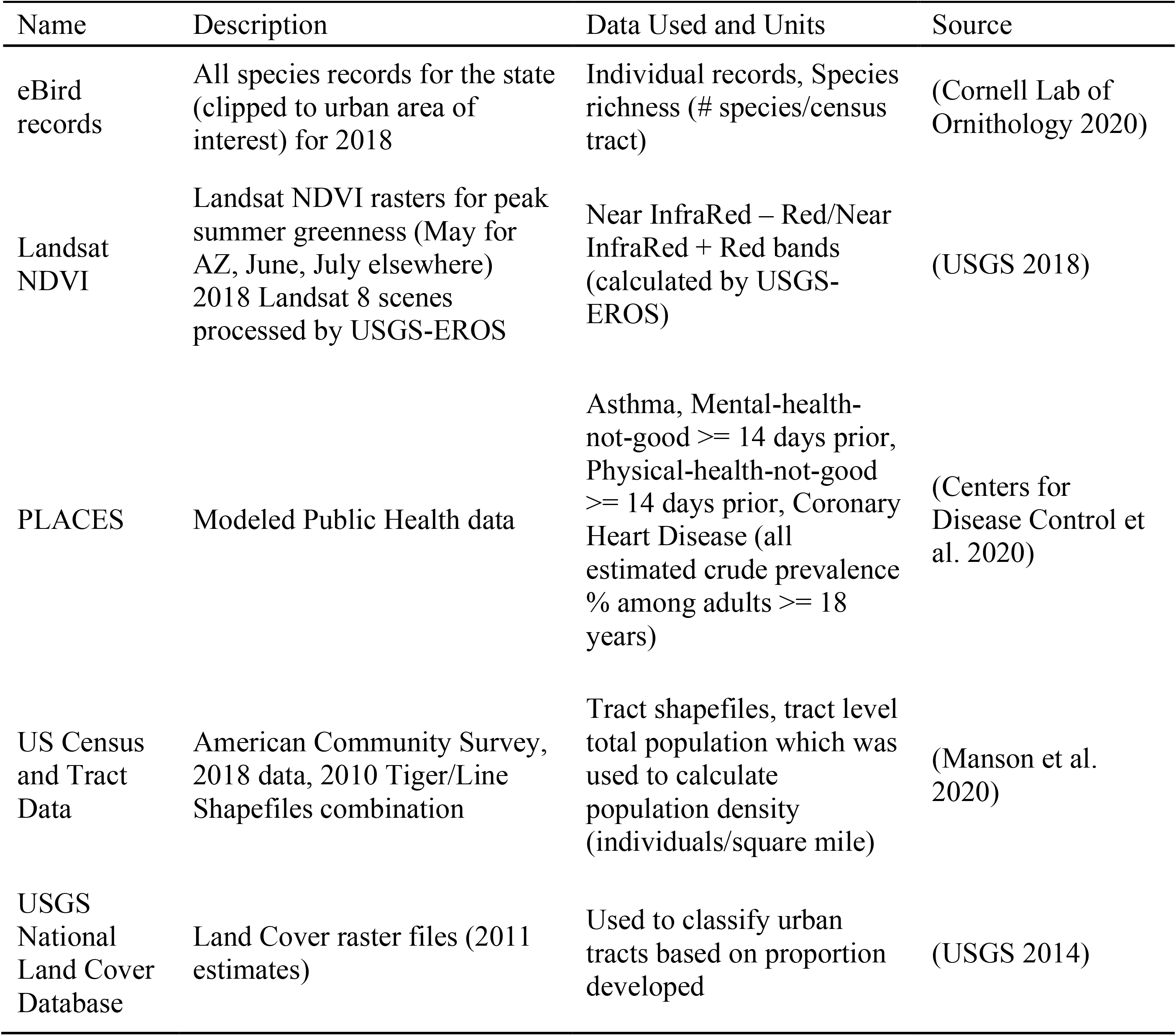
Data descriptions and sources.

## Results

While eBird richness correlated with several public health outcomes and with NDVI, it did not, overall, appear to be a proxy for greenness. In fact, neither richness nor greenness related to the four public health outcomes to a degree that would justify their use as predictors due to wide scatter and apparent (unmeasured) differences among metro areas. When combining census tract data across all three metro areas combined, eBird record species richness has a weak positive correlation with NDVI (R_s_ = 0.25, Table 2, Fig. 1) and is weakly negatively correlated with human population density and the aerial proportion developed (R_s_ = -0.44, -0.30, respectively, Table 2, Fig. 1). When aggregated across metro areas, there are weak negative relationships between species richness and current adult asthma, self-assessed poor mental health, and self-assessed poor physical health (R_s_ = -0.38, -0.43, -.38 respectively, Table 2) whereas NDVI is less correlated with these three health outcomes. However, relationships among the variables differed depending on the metro area (Tables 3, 4, 5, Figs. 2-4).

**Table 2.** Spearman’s correlation coefficients among urban development, bird species richness and public health variables for combined Phoenix, AZ, Kansas City, MO and Albany, NY census tracts. n = 1199

|  | Pop. Dens. | NDVI | Richness | Developed | Asthma | Mental Health | Phys Health |
| --- | --- | --- | --- | --- | --- | --- | --- |
| Pop. Dens. |  |  |  |  |  |  |  |
| NDVI | -0.36*** |  |  |  |  |  |  |
| Richness | -0.44*** | 0.25*** |  |  |  |  |  |
| Developed | 0.62*** | -0.24*** | -0.30*** |  |  |  |  |
| Asthma | 0.40*** | -0.19*** | -0.38*** | 0.20*** |  |  |  |
| Mental Health | 0.43*** | -0.18*** | -0.43*** | 0.25*** | 0.86*** |  |  |
| Phys Health | 0.27*** | -0.25*** | -0.38*** | 0.30*** | 0.70*** | 0.70*** |  |
| CHD | -0.08** | 0.09** | -0.06 | 0.15*** | 0.33*** | 0.17*** | 0.72*** |

\*, \*\* and \*\*\* indicate $p < 0.05$ , $p < 0.01$ and $p < 0.001$ , respectively. Pop. Dens. = human population density (individuals $\text{mi.}^{-2}$ ), NDVI = Landsat 8 Normalized Difference Vegetation Index ( $\text{NIR-R}/\text{NIR+R}$ ), Richness = of spp. recorded in eBird records for 2018, Developed = aerial proportion of census tract mapped as developed by USGS NLCD. Asthma, Mental Health, Phys Health, CHD estimated as crude prevalence (%).

**Figure 1.**
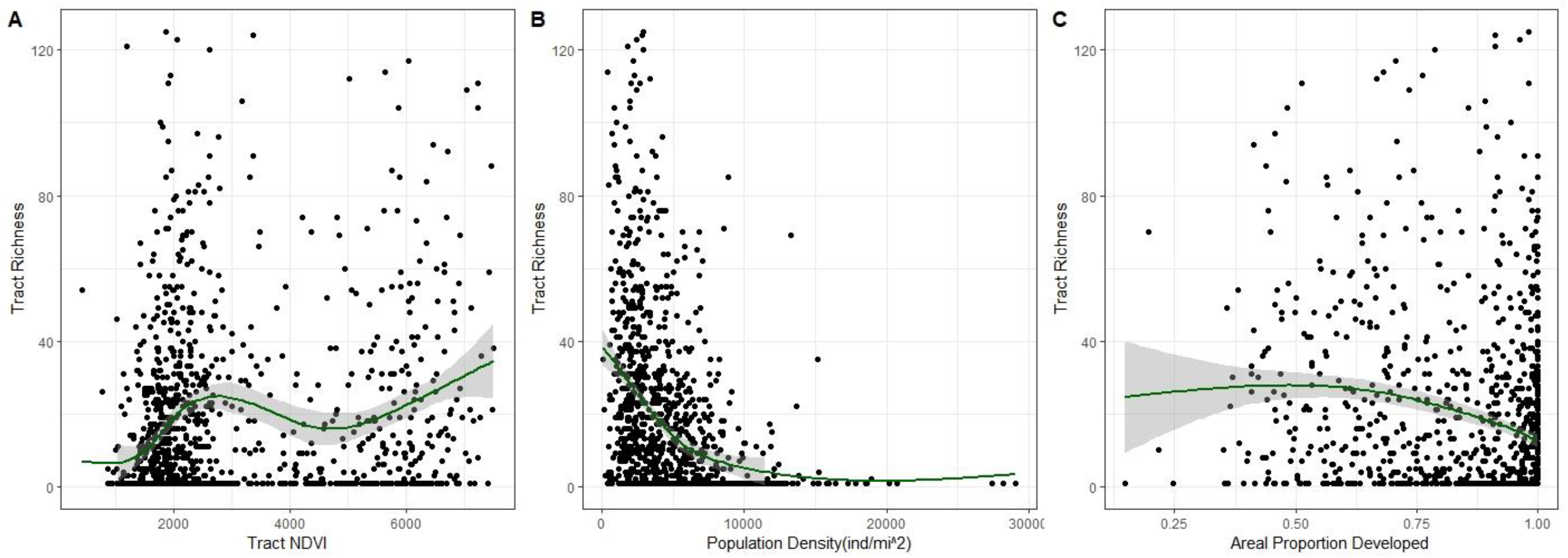
Relationships between urbanization variables and eBird record species richness with loess curves for census tracts combined across the metro areas of Phoenix, AZ, Kansas City, MO, and Albany, NY. eBird record species richness was quantified as the number of unique species included in 2018 eBird records for a given census tract. A) NDVI (Landsat 8 Normalized Difference Vegetation Index (NIR-R/NIR+R), summer, 2018), B) Human Population Density (individuals mi^-2^), and C) Aerial proportion of the census tract that is developed based on USGS-National Land Cover Database raster data. The y-axis is truncated at 120 species to improve visualization; this does not affect the loess line. N=1199.

**Table 3.** Spearman’s correlation coefficients among urban development, bird species richness and public health variables for Phoenix, AZ metro census tracts. n = 852

|  | Pop. Dens. | NDVI | Richness | Developed | Asthma | Mental Health | Phys Health |
| --- | --- | --- | --- | --- | --- | --- | --- |
| Pop. Dens. |  |  |  |  |  |  |  |
| NDVI | -0.27*** |  |  |  |  |  |  |
| Richness | -0.47*** | 0.33*** |  |  |  |  |  |
| Developed | 0.58*** | -0.17*** | -0.25*** |  |  |  |  |
| Asthma | 0.53*** | -0.52*** | -0.44*** | 0.29*** |  |  |  |
| Mental Health | 0.55*** | -0.51*** | -0.44*** | 0.28*** | 0.95*** |  |  |
| Phys Health | 0.35*** | -0.46*** | -0.36*** | 0.32*** | 0.77*** | 0.65*** |  |
| CHD | 0.02 | -0.19*** | -0.06 | 0.25*** | 0.25*** | 0.06 | 0.73*** |

\*, \*\* and \*\*\* indicate $p < 0.05$ , $p < 0.01$ and $p < 0.001$ , respectively. Pop. Dens. = human population density (individuals $\text{mi.}^{-2}$ ), NDVI = Landsat 8 Normalized Difference Vegetation Index ( $\text{NIR-R}/\text{NIR+R}$ ), Richness = of spp. recorded in eBird records for 2018, Developed = aerial proportion of census tract mapped as developed by USGS NLCD. Asthma, Mental Health, Phys Health, CHD estimated as crude prevalence (%).

**Table 4.**
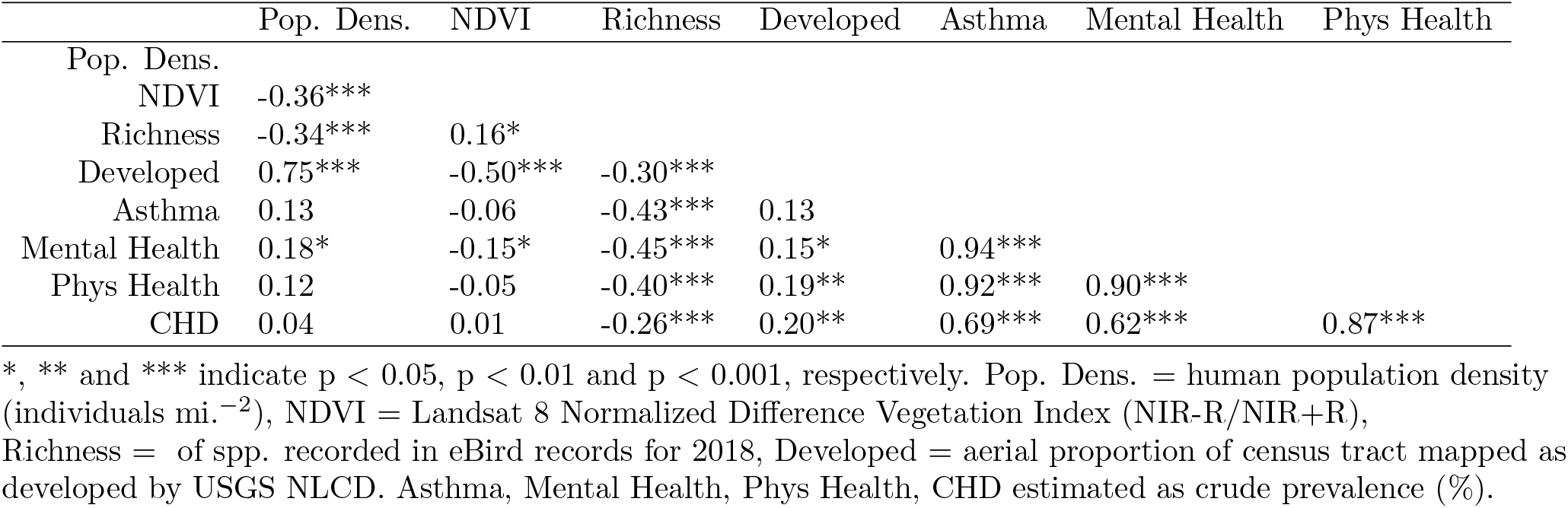
Spearman’s correlation coefficients among urban development, bird species richness and public health variables for Kansas City, MO metro census tracts. n = 224

**Table 5.** Spearman’s correlation coefficients among urban development, bird species richness and public health variables for Albany, NY metro census tracts. n = 123

|  | Pop. Dens. | NDVI | Richness | Developed | Asthma | Mental Health | Phys Health |
| --- | --- | --- | --- | --- | --- | --- | --- |
| Pop. Dens. |  |  |  |  |  |  |  |
| NDVI | -0.59*** |  |  |  |  |  |  |
| Richness | -0.57*** | 0.43*** |  |  |  |  |  |
| Developed | 0.83*** | -0.65*** | -0.57*** |  |  |  |  |
| Asthma | 0.63*** | -0.65*** | -0.47*** | 0.55*** |  |  |  |
| Mental Health | 0.68*** | -0.67*** | -0.48*** | 0.57*** | 0.97*** |  |  |
| Phys Health | 0.34*** | -0.45*** | -0.36*** | 0.28** | 0.83*** | 0.75*** |  |
| CHD | 0.01 | -0.22* | -0.11 | 0.07 | 0.41*** | 0.30*** | 0.77*** |

\*, \*\* and \*\*\* indicate $p < 0.05$ , $p < 0.01$ and $p < 0.001$ , respectively. Pop. Dens. = human population density (individuals $\text{mi.}^{-2}$ ), NDVI = Landsat 8 Normalized Difference Vegetation Index ( $\text{NIR-R}/\text{NIR+R}$ ), Richness = of spp. recorded in eBird records for 2018, Developed = aerial proportion of census tract mapped as developed by USGS NLCD. Asthma, Mental Health, Phys Health, CHD estimated as crude prevalence (%).

**Figure 2.**
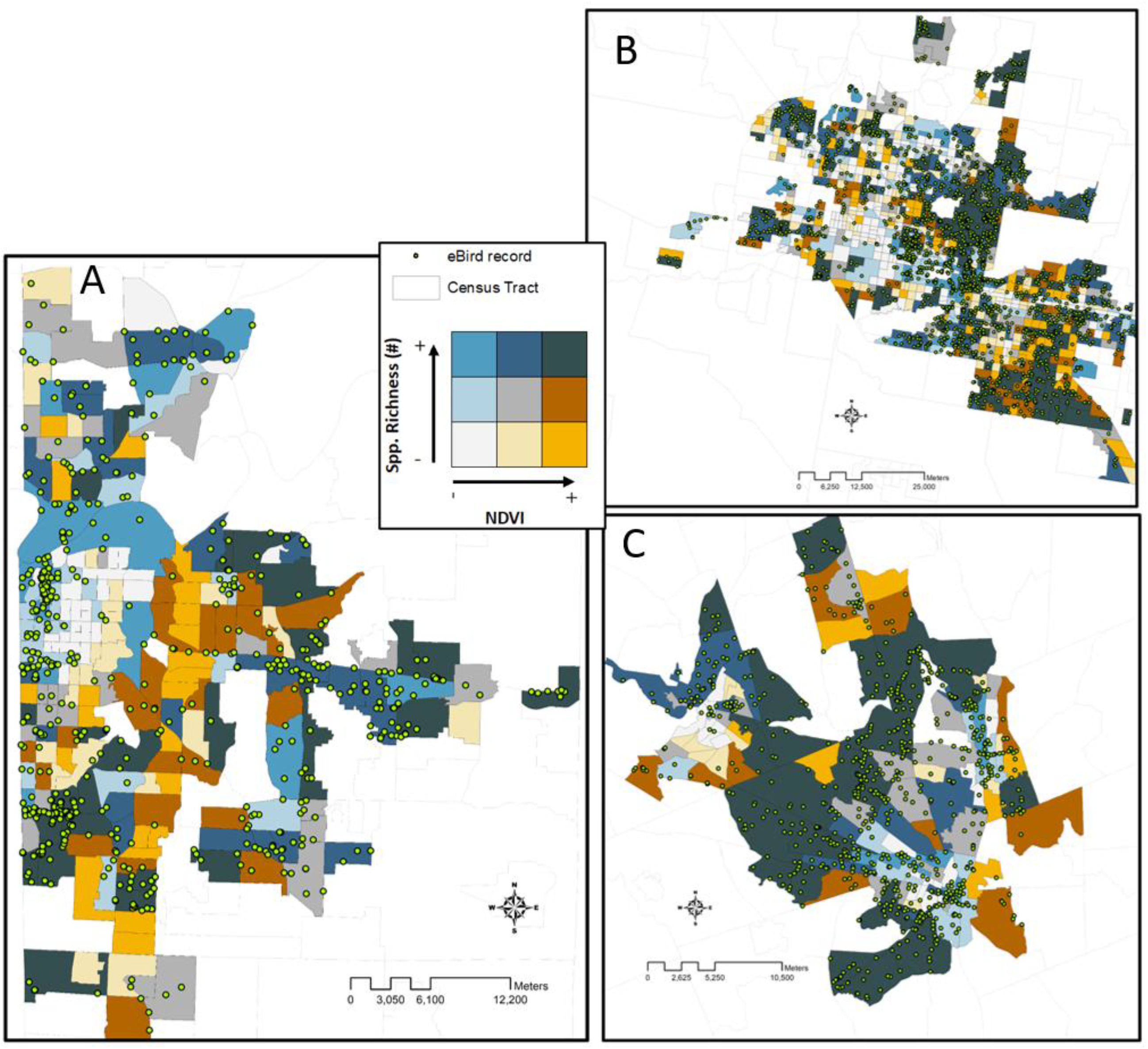
2018 eBird records, eBird species richness and summer 2018 greenness (Landsat 8 Normalized Difference Vegetation Index (NIR-R/NIR+R)) in the urban census tracts of the Kansas City, MO (A), Phoenix, AZ (B) and Albany, NY metro areas.

**Figure 3.**
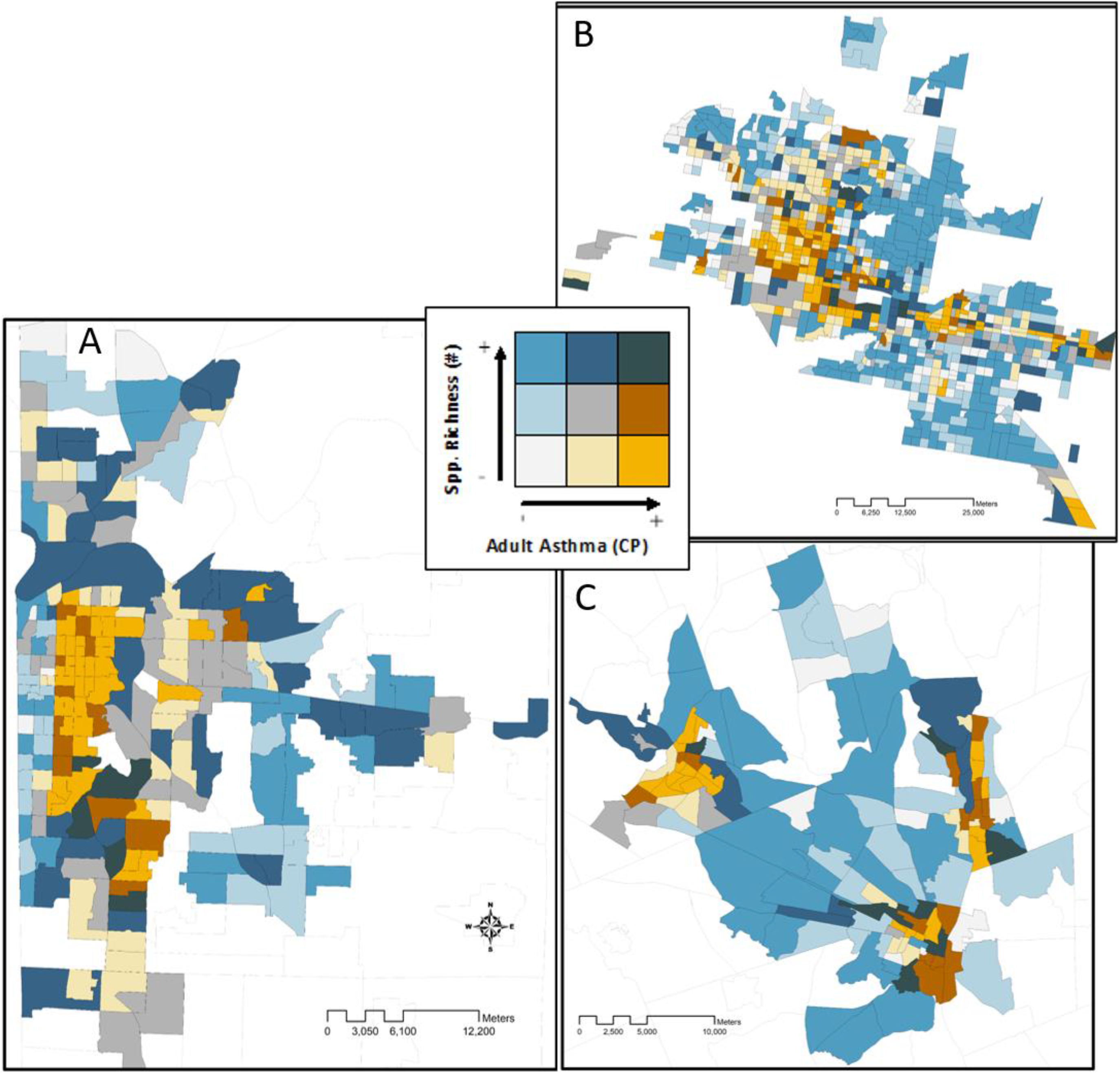
eBird record species richness (2018) and currently active adult asthma (crude prevalence, %) in the urban census tracts of the Kansas City, MO (A), Phoenix, AZ (B) and Albany, NY metro areas.

**Figure 4.**
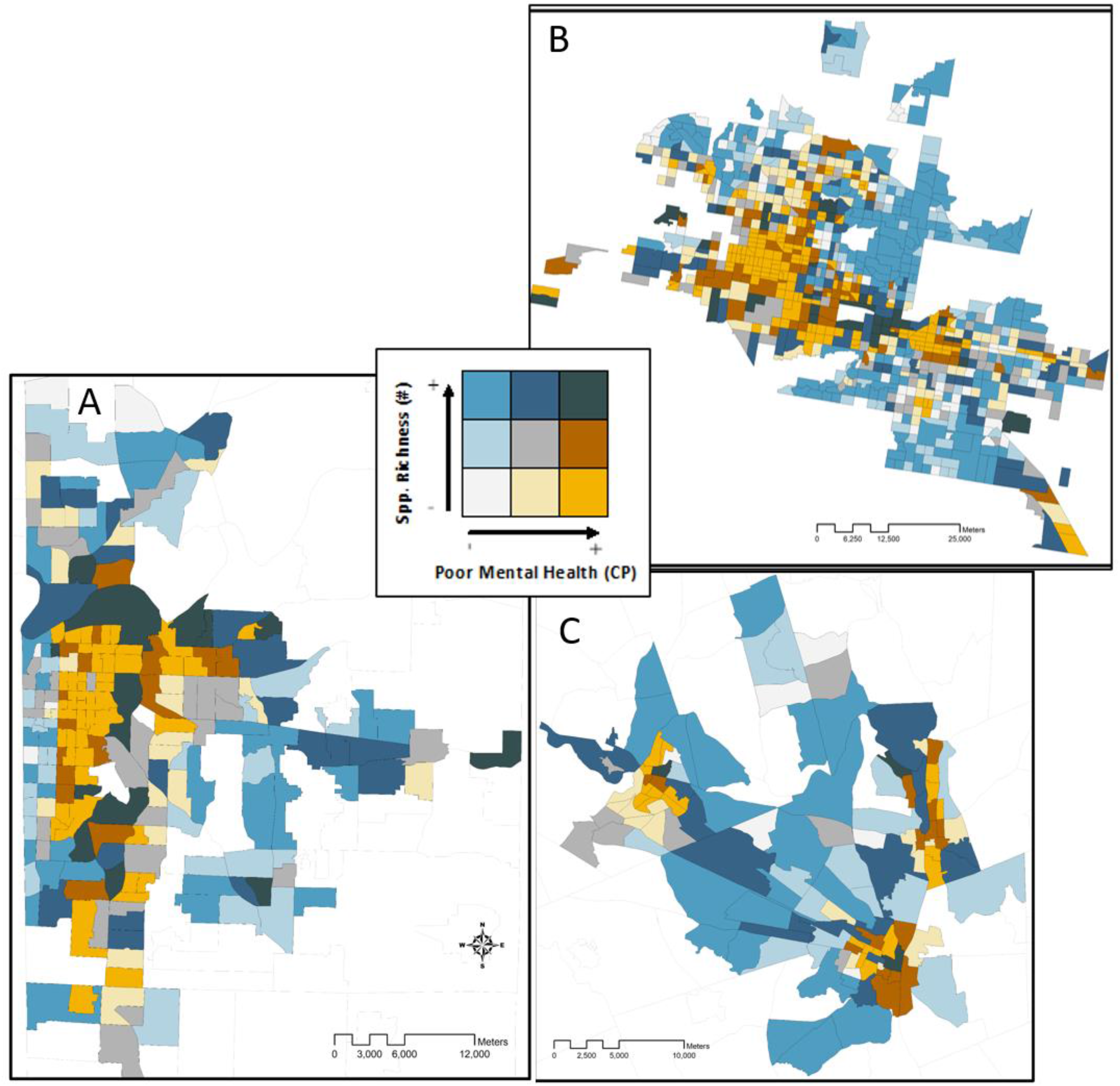
eBird record species richness (2018) and self-assessed poor mental health in the previous 14 days (crude prevalence, %) in the urban census tracts of the Kansas City, MO (A), Phoenix, AZ (B) and Albany, NY metro areas.

While NDVI was positively correlated with species richness in all three metro areas, the correlation was weaker in Kansas City (R_s_ = 0.16, Table 4, Fig. S1, S2) than in Phoenix (R_s_ = 0.33, Table 3, Fig. S3, S4) and Albany (R_s_ = 0.43, Table 5, Fig. S5, S6). Species richness decreased with increasing population density and development in all three metro areas, but the relationship was strongest in the Albany, NY area (Tables 3, 4, 5, Fig. 2, S1, S3, S5). Associations between species richness and the four health outcomes also differed among the metro areas. Species richness was more strongly correlated with asthma (R_s_ = -0.43), mental health (R_s_ = -0.45) and physical health (R_s_ = -0.40) than NDVI was (R_s_ = -0.06 ns, R_s_ = -0.15, R_s_ = -0.05 ns, respectively) in Kansas City (Table 4, Fig. 3, 4, S2); however, the NDVI was more correlated with those health outcomes than species richness in Phoenix (Table 3, Fig. 3, 4, S4) and the Albany area, NY (Table 5, Fig. 3, 4, S6). Chronic heart disease did not correlate with or is only very weakly correlated with any of the urbanization, greenness or eBird variables, regardless of metro area (Tables 2, 3, 4, 5).

## Discussion

My objective was to determine whether eBird data might be an improved estimator of nature access over simple greenness indices. While eBird species richness was related to greenness, measures of urbanization and several human health factors, the correlations varied by metro area and were not always strong. eBird data was a clearer correlate with health factors than NDVI in one metro area (Kansas City) but was not an improvement over NDVI in for Phoenix or the Albany area of NY. However, the spatial relationship between asthma and mental health and bird species richness was striking. Both asthma and poor mental health increased as bird diversity and NDVI decreased such that city centers and areas of high urbanization are readily apparent on bivariate choropleth maps (Fig. 3, 4). Despite the weakness of some relationships, eBird and other volunteer gathered data could be useful covariates when modeling access to nature and biodiversity and implications for public health, particularly if some limiting factors were addressed. One should consider the ecological and cultural situation of the population center, biases associated with volunteer collected data, potential confounders including socio-economic status, and limitations on data precision.

Differences in relationships depending on the metro area and location should not be discounted-blanket statements regarding eBird data or greenness and public health based on single location studies should be avoided (Fuertes et al. 2014). For example, greenness in the Phoenix area is concentrated in ephemeral waterways and in heavily populated and irrigated areas whereas greenness has a completely different distribution in upstate New York-a much more humid climate. Bird population distributions and whether the metro area is part of a migratory fly-zone should also be considered. Further, cultural differences within and across cities may influence birding and contribution activities and public values placed on parks and other open spaces.

Beyond considering the location of the investigation, controlling for other variables that may confound the relationships among NDVI, eBird record species richness and public health outcomes might make eBird and other volunteer gathered biodiversity data more useful. Citizen science data are vulnerable to bias since observation records depend on the knowledge, geographic location, biases, and effort of the contributor. Data such as these are typically concentrated around population centers since most citizen scientists are associated with urban, suburban and exurban areas (Zhang 2020). I attempted to control for this bias by limiting my investigation to only ‘urban’ census tracts with population densities > 1500 people/square mile and at least 40% development. However, variables such as effort and knowledge of the birder were not controlled for in this study, which may have biased the results. Socioeconomic factors should also be controlled for when attempting to model the relationships between nature access and human health. Often, green space, healthy food access and access to medical services are all limited in neighborhoods with lower socioeconomic status, implying that while green space may be a correlate to poor health, the relationship may not be causal (Jarvis et al. 2020). I did not control for SES in this study; however, the PLACES data does include an estimate of health insurance access which may serve as a proxy for SES (Centers for Disease Control et al. 2020) that one could investigate in the future. Other factors also interact with health outcomes; while asthma and allergy prevalence has been related to the proximity of green space (James et al. 2015, Shanahan et al. 2015, Sbihi et al. 2019), asthma may also be related to air pollution, which tends to be higher in urbanized areas.

Limitations in the spatial and numerical precision of the data may also have influenced the results of this investigation. A census tract can have a large or small spatial footprint depending on population density. When one aggregates fine scale data such as NDVI rasters or eBird points to larger scale polygons such as census tract, data, details, and nuance are lost or compressed with loss. In addition, human health data provided in the PLACES data is modeled and interpolated, further limiting precision and impinging relationship discernment. Therefore, the spatial coarseness of the public health data and confounders associated with eBird sightings may limit their utility as a predictor of public health measures. Finer scale public health data would alleviate this limitation. In the interest of expediency, I limited my dataset to greenness and eBird data from only one year. An investigation across years would be informative and might enhance the value of eBird and other volunteer collected data toward understanding relationships between access to nature and public health outcomes.

Despite the limitations and potential confounders, eBird data does appear to have some value in understanding the relationships among proximity to greenness, biodiversity, and human health measures.

## Conclusion

Greenness indices are often used as a proxy for access to nature when studying the relationship between nature and public health; however, greenness indices do not provide measure of the “quality” of the proximate nature experience. One possible remedy for this oversight is the use of citizen science biodiversity data, such as eBird records, to relate greenness and species richness-a measure of habitat quality. In this study, I found that eBird record richness was correlated with measures of greenness, urban development, and public health outcome estimates in all three metro areas investigated; however, the variability of the data and potential confounders do not recommend eBird data as an improved estimator of nature access above and beyond simple greenness indices. However, eBird and other volunteer collected biodiversity data could inform models as a covariate.

## Data Availability

All data are publicly available via the CDC website, USGS, US Census and Cornell lab of Ornithology (upon request)

https://ebird.org/science/use-ebird-data/download-ebird-data-products

https://nrcs.app.box.com/v/gateway/folder/2222277200

https://chronicdata.cdc.gov/500-Cities-Places/PLACES-Census-Tract-Data-GIS-Friendly-Format-2020-/yjkw-uj5s

https://www.nhgis.org/

## References

Buckley, A. 2015. Making Bivariate Choropleth Maps with ArcMap. ArcGIS Blog. esri.

Centers for Disease Control, Robert Wood Johnson Foundation, and CDC Foundation. 2020. PLACES: Local Data for Better Health.in CDC, editor. CDC, https://www.cdc.gov/places/index.html.

Cornell Lab of Ornithology. 2020. eBird Basic Dataset. EBD_relDec-2020, Ithaca, NY.

Esri. 2017. ArcGIS Desktop 10.6.1.

Fuertes, E., I. Markevych, A. von Berg, C. P. Bauer, D. Berdel, S. Koletzko, D. Sugiri, and J. Heinrich. 2014. Greenness and allergies: evidence of differential associations in two areas in Germany. Journal of Epidemiology and Community Health 68:787–790.

Fuller, R. A., K. N. Irvine, P. Devine-Wright, P. H. Warren, and K. J. Gaston. 2007. Psychological benefits of greenspace increase with biodiversity. Biology letters 3:390–394.

Hanski, I., L. von Hertzen, N. Fyhrquist, K. Koskinen, K. Torppa, T. Laatikainen, P. Karisola, P. Auvinen, L. Paulin, M. J. Makela, E. Vartiainen, T. U. Kosunen, H. Alenius, and T. Haahtela. 2012. Environmental biodiversity, human microbiota, and allergy are interrelated. Proceedings of the National Academy of Sciences of the United States of America 109:8334–8339.

James, P., R. F. Banay, J. E. Hart, and F. Laden. 2015. A review of the health benefits of greenness. Current epidemiology reports 2:131–142.

Jarvis, I., S. Gergel, M. Koehoorn, and M. van den Bosch. 2020. Greenspace access does not correspond to nature exposure: Measures of urban natural space with implications for health research. Landscape and Urban Planning 194:103686.

Jennings, V., L. Larson, and J. Yun. 2016. Advancing sustainability through urban green space: Cultural ecosystem services, equity, and social determinants of health. International Journal of environmental research and public health 13:196.

Leveau, L. M., F. I. Isla, and M. I. Bellocq. 2020. From town to town: Predicting the taxonomic, functional and phylogenetic diversity of birds using NDVI. Ecological Indicators 119:11.

Manson, S., J. Schroeder, D. Van Riper, T. Kugler, and S. Ruggles. 2020. IPUMS National Historical Geographic Information System: Version 15.0 [dataset]. in IPUMS, editor. IPUMS, Minneapolis, MN.

Markevych, I., J. Schoierer, T. Hartig, A. Chudnovsky, P. Hystad, A. M. Dzhambov, S. de Vries, M. Triguero-Mas, M. Brauer, M. J. Nieuwenhuijsen, G. Lupp, E. A. Richardson, T. Astell-Burt, D. Dimitrova, X. Q. Feng, M. Sadeh, M. Standl, J. Heinrich, and E. Fuertes. 2017. Exploring pathways linking greenspace to health: Theoretical and methodological guidance. Environmental Research 158:301–317.

Mavoa, S., M. Davern, M. Breed, and A. Hahs. 2019. Higher levels of greenness and biodiversity associate with greater subjective wellbeing in adults living in Melbourne, Australia. Health & place 57:321–329.

Melles, S. J. 2005. Urban bird diversity as an indicator of human social diversity and economic inequality in Vancouver, British Columbia. Urban habitats 3:25–48.

Mills, J. G., A. Bissett, N. J. C. Gellie, A. J. Lowe, C. A. Selway, T. Thomas, P. Weinstein, L. S. Weyrich, and M. F. Breed. 2020. Revegetation of urban green space rewilds soil microbiotas with implications for human health and urban design. Restoration Ecology 28:S322–S334.

R. Core Team. 2020. R: A language and environment for statistical computing. Vienna, Austria.

Reid, C. E., L. D. Kubzansky, J. Li, J. L. Shmool, and J. E. Clougherty. 2018. It’s not easy assessing greenness: a comparison of NDVI datasets and neighborhood types and their associations with self-rated health in New York City. Health & place 54:92–101.

Sbihi, H., R. C. Boutin, C. Cutler, M. Suen, B. B. Finlay, and S. E. Turvey. 2019. Thinking bigger: How early-life environmental exposures shape the gut microbiome and influence the development of asthma and allergic disease. Allergy 74:2103–2115.

Shanahan, D. F., R. A. Fuller, R. Bush, B. B. Lin, and K. J. Gaston. 2015. The health benefits of urban nature: how much do we need? Bioscience 65:476–485.

Tratalos, J., R. A. Fuller, K. L. Evans, R. G. Davies, S. E. Newson, J. J. Greenwood, and K. J. Gaston. 2007. Bird densities are associated with household densities. Global Change Biology 13:1685–1695.

USGS. 2014. NLCD 2011 Land Cover (2011 Edition). USGS, Sioux Falls, SD..

USGS. 2018. USGS EROS Archive - Vegetation Monitoring - Landsat 8.

Wang, R., and J. A. Gamon. 2019. Remote sensing of terrestrial plant biodiversity. Remote Sensing of Environment 231:111218.

Zhang, G. 2020. Spatial and Temporal Patterns in Volunteer Data Contribution Activities: A Case Study of eBird. ISPRS International Journal of Geo-Information 9:597.

