## Supplemental Figures 1-6 for "Citizen Science and Public Health- Can eBird data inform relationships between public health and access to biodiversity?"

Supplement

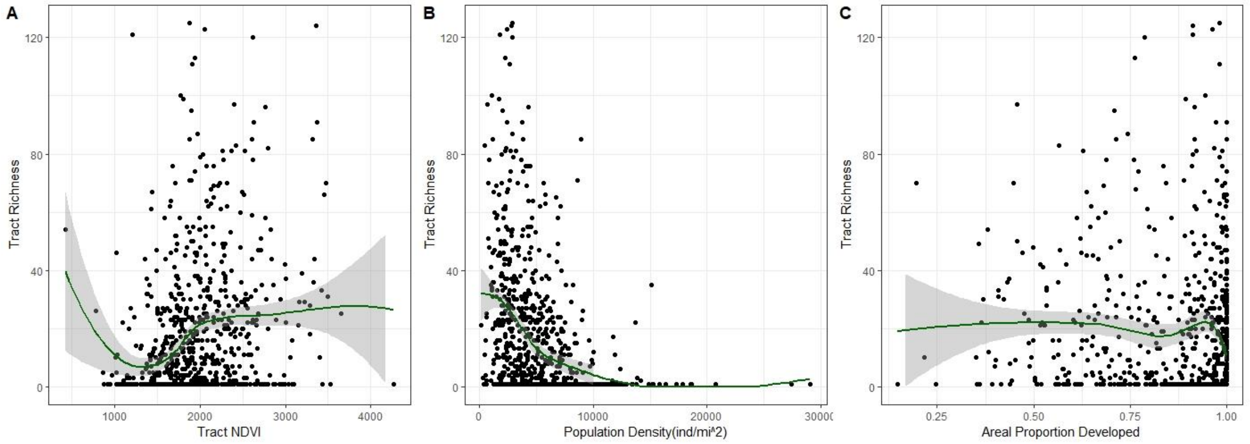


Figure S1. Relationships between urbanization variables and eBird record species richness with loess curves for urban census tracts in the Kansas City, MO metro area. eBird record species richness was quantified as the number of unique species included in 2018 eBird records for a given census tract. A) NDVI (Landsat 8 Normalized Difference Vegetation Index (NIR-R/NIR+R), summer, 2018), B) Human Population Density (individuals mi$^{-2}$), and C) Aerial proportion of the census tract that is developed based on USGS-National Land Cover Database raster data. The y-axis is truncated at 120 species to improve visualization; this does not affect the loess line. N=224.

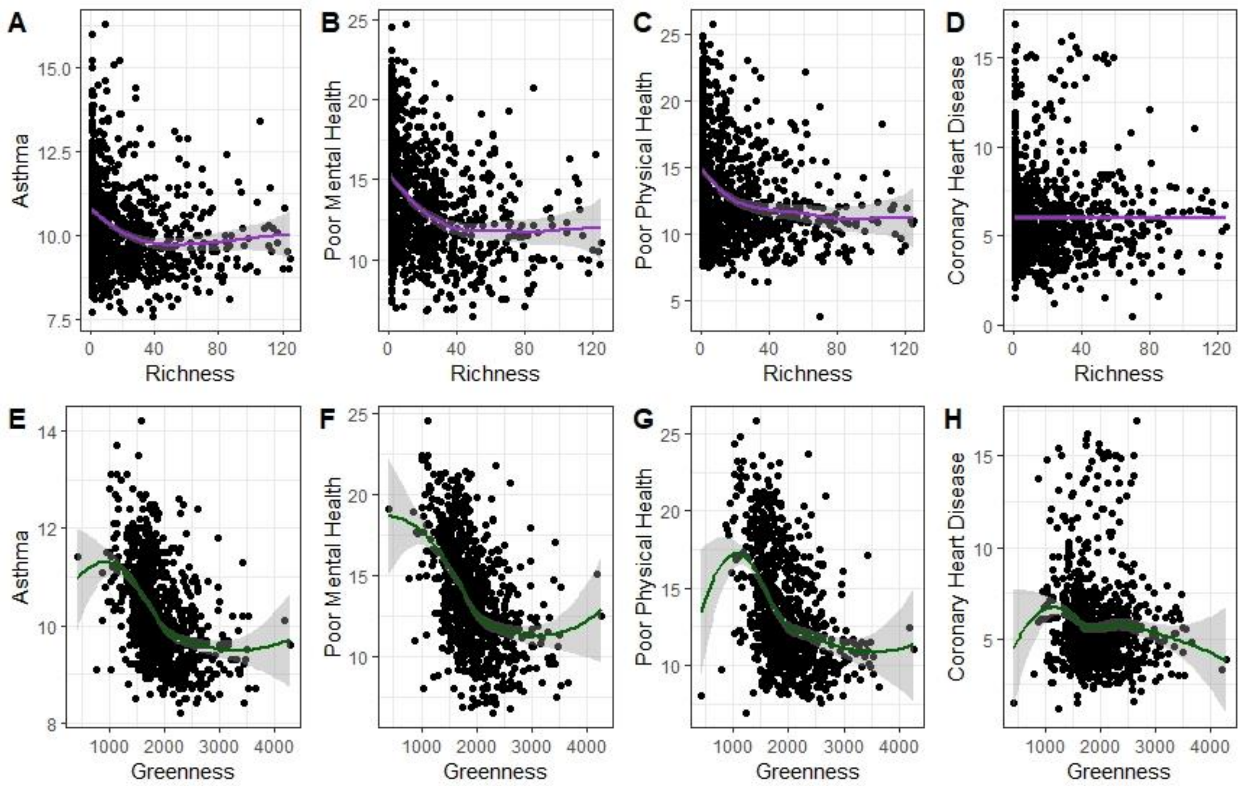


Figure S2. Relationships between public health variables and eBird record species richness (A, B, C, D) or NDVI (E, F, G, H) with loess curves for urban census tracts in the Kansas City, MO metro area. eBird record species richness was quantified as the number of unique species included in 2018 eBird records for a given census tract; NDVI data are from summer 2018 and are calculated as NIR-R/NIR+R. All public health variables are estimated crude prevalence and are A) Current adult asthma, B) Self- assessed poor mental health within the last 14 days, C) Self- assessed poor mental health within the last 14 days, and D) coronary heart disease. The x-axis is truncated at 120 species to improve visualization; this does not affect the loess line. N=224.

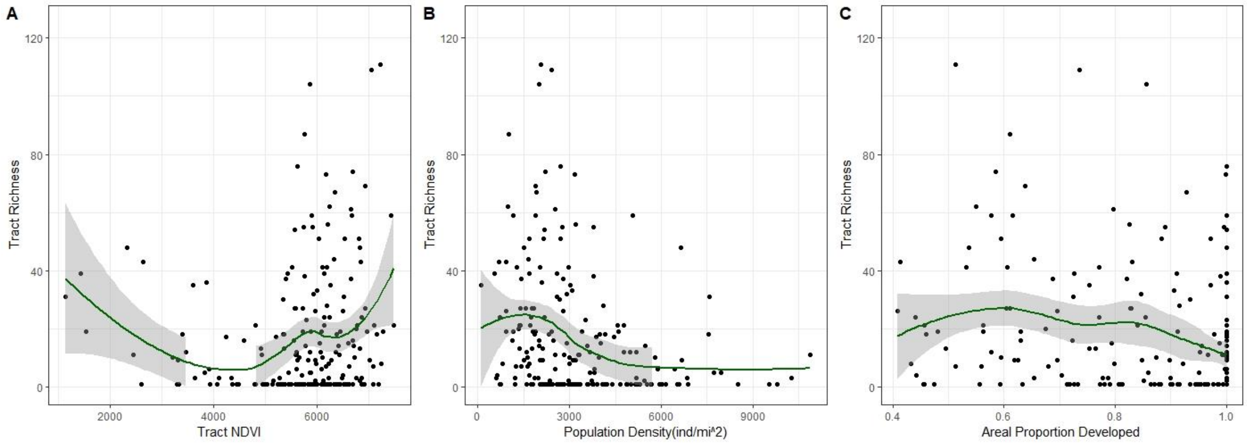


Figure S3. Relationships between urbanization variables and eBird record species richness with loess curves for urban census tracts in the Phoenix, AZ metro area. eBird record species richness was quantified as the number of unique species included in 2018 eBird records for a given census tract. A) NDVI (Landsat 8 Normalized Difference Vegetation Index (NIR-R/NIR+R), summer, 2018), B) Human Population Density (individuals $mi^{-2}$), and C) Aerial proportion of the census tract that is developed based on USGS-National Land Cover Database raster data. The y-axis is truncated at 120 species to improve visualization; this does not affect the loess line. N=852.

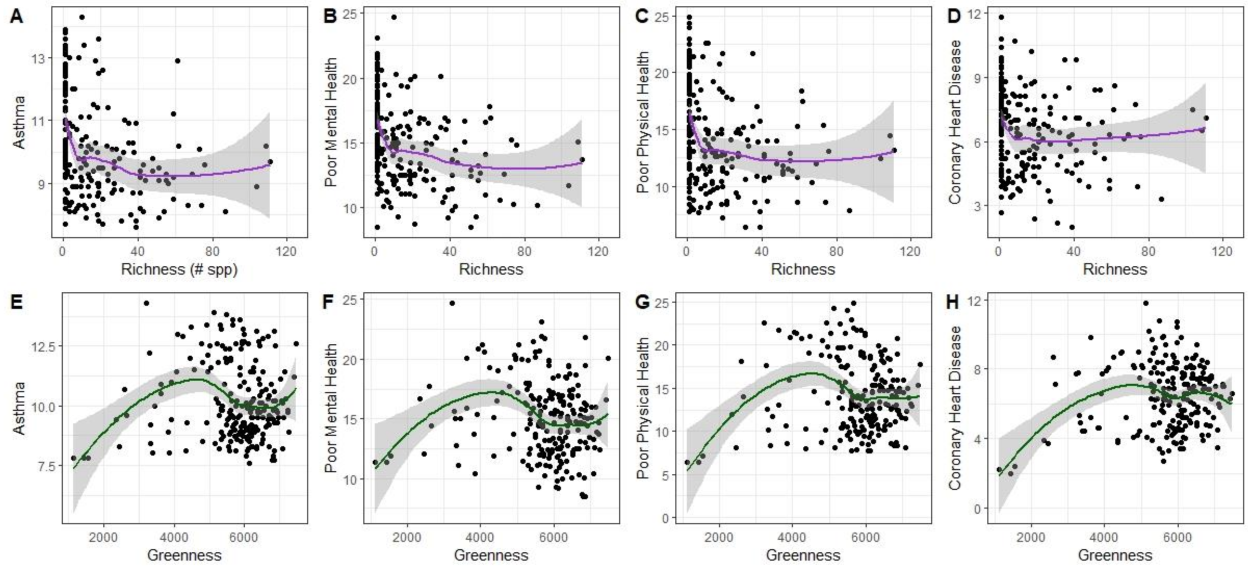


Figure S4. Relationships between public health variables and eBird record species richness (A, B, C, D) or NDVI (E, F, G, H) with loess curves for urban census tracts in the Phoenix, AZ metro area. eBird record species richness was quantified as the number of unique species included in 2018 eBird records for a given census tract; NDVI data are from summer 2018 and are calculated as NIR-R/NIR+R. All public health variables are estimated crude prevalence and are A) Current adult asthma, B) Self- assessed poor mental health within the last 14 days, C) Self- assessed poor mental health within the last 14 days, and D) coronary heart disease. The x-axis is truncated at 120 species to improve visualization; this does not affect the loess line. N=852.

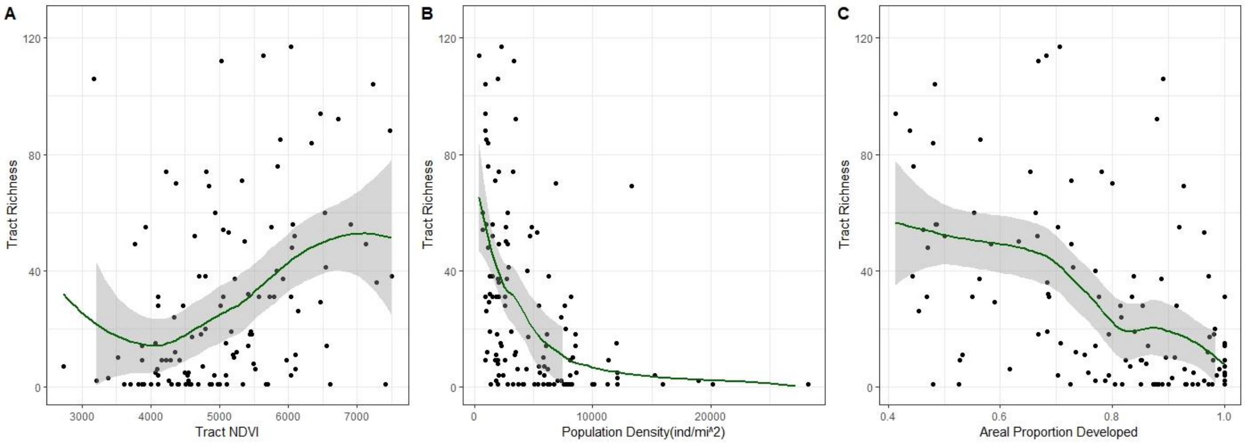


Figure S5. Relationships between urbanization variables and eBird record species richness with loess curves for urban census tracts in the Albany area, NY. eBird record species richness was quantified as the number of unique species included in 2018 eBird records for a given census tract. A) NDVI (Landsat 8 Normalized Difference Vegetation Index (NIR-R/NIR+R), summer, 2018), B) Human Population Density (individuals $mi^{-2}$), and C) Aerial proportion of the census tract that is developed based on USGS-National Land Cover Database raster data. The y-axis is truncated at 120 species to improve visualization; this does not affect the loess line. N=123.

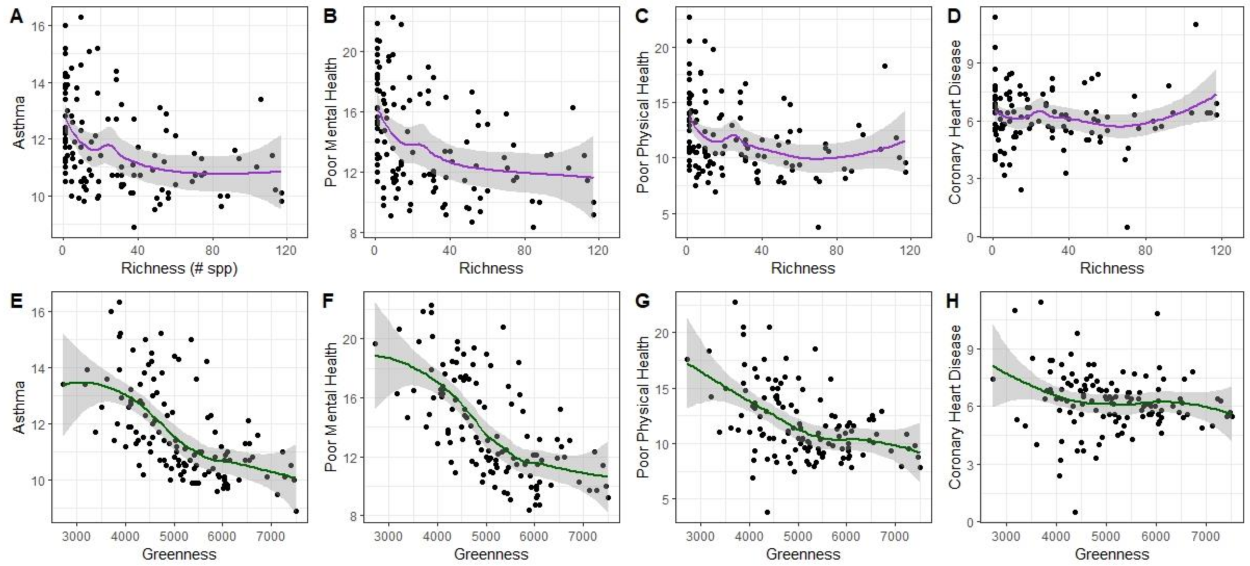


Figure S6. Relationships between public health variables and eBird record species richness (A, B, C, D) or NDVI (E, F, G, H) with loess curves for urban census tracts in the Albany area, NY. eBird record species richness was quantified as the number of unique species included in 2018 eBird records for a given census tract; NDVI data are from summer 2018 and are calculated as NIR-R/NIR+R. All public health variables are estimated crude prevalence and are A) Current adult asthma, B) Self- assessed poor mental health within the last 14 days, C) Self- assessed poor mental health within the last 14 days, and D) coronary heart disease. The x-axis is truncated at 120 species to improve visualization; this does not affect the loess line. N=123.
